# THE *MUC5B* PROMOTOR POLYMORPHISM ASSOCIATES WITH SEVERE COVID-19

**DOI:** 10.1101/2020.05.12.20099333

**Authors:** Coline H.M. van Moorsel, Joanne J. van der Vis, Claudia Benschop, Henk J.T. Ruven, Marian Quanjel, Jan C. Grutters

## Abstract

**Background:** Diversity in response to exposition to severe acute respiratory syndrome coronavirus 2 (SARS-CoV-2) is common and may be related to the innate immune response. The mucin MUC5B is an important component of the innate immune response and expression levels are associated with the *MUC5B* promoter polymorphism, rs35705950. The high expressing T-allele of rs35705950 is an accepted risk allele for a non-infectious aging lung disease called idiopathic pulmonary fibrosis (IPF). However, given the theory of trade-offs in aging lung disease and the importance of high expression for an adequate immune response, we hypothesize that the T-allele is protective against severe coronavirus disease 2019 (COVID-19).

**Methods:** We collected demographics, radiology, survival data and *MUC5B* rs35705950 allele status for 108 patients requiring hospitalisation for COVID-19 at St Antonius Hospital in The Netherlands. For comparison of allele frequencies and allele carriership with a white control cohort, the patient cohort was divided in a white (n=83) and non-white cohort.

**Results:** The patients had a median age of 66 years and consisted predominantly of males (74%) and 23 patients (21 %) died. The T-allele frequencies of rs35705950 in white patients was 0.04 which was significantly lower than the T-allele frequency of 0.10 in white controls (p= 0.02). Moreover, comparison of the number of carriers and non-carriers of the T allele showed that only 8.4% of patients carried the T-allele versus 18% of controls (p=0.029; OR= 0.41, CI=0.19-0.94).

**Conclusions:** The *MUC5B* rs35705950 promoter polymorphism associates with COVID-19. The risk allele (T) for IPF is protective against the development of severe COVID-19 disease. This is a further example of a trade-off between optimal expression levels in the respiratory system which associates with aging diseases. However, these results require further investigation.

## Introduction

The current coronavirus disease COVID-19 pandemic illustrates the diversity in response to exposition to severe acute respiratory syndrome coronavirus 2 (SARS-CoV-2). Response to infection ranges between asymptomatic and death from organ failure, of which the latter is most commonly observed in the elderly^1^. Such differences are associated with aging but may also be influenced by the genetic constitution of the host. However, to date genetic predisposing factors to COVID-19 are unknown.

Diversity in response to exposition to SARS-CoV-2 may be related to host factors associated with airway defense. The gel-forming mucin 5B (MUC5B) is part of the mucus that covers the surface of the respiratory epithelium and forms the first line of defense against respiratory pathogens.^2,3,4^. In vivo studies in mice showed that presence of Muc5B is essential for mucociliary clearance, a mechanism crucial for proper airway defense ^5^. Mice heterozygous for a knock-out allele of Muc5B secreted approximately 50% of the wild-type level of Muc5b and demonstrate reduced mucociliary clearance ^5^. Decreased mucociliary clearance is associated with aging in humans ^6^ and with aging in mice ^7^. Furthermore, aged mice with reduced mucociliary clearance had significantly reduced Muc5b levels in comparison with young mice ^7^. Constitutive expression levels of MUC5B are associated with a common promoter polymorphism, rs35705950 of the encoding gene *MUC5B*. The minor rs35705950 T allele is associated with high expression levels of MUC5B and the major G allele is associated with low expression levels ^8,9^. The high expressing T-allele is a known risk factor for idiopathic pulmonary fibrosis (IPF)^8^, a fatal aging lung disease of unknown cause predominately affecting older males with a history of smoking. IPF is a non-infectious disease of the distal lung, caused by damage of the alveolar epithelium following by progressive fibrogenesis ^10^.

Recently it was shown that aging lung diseases such as IPF and COPD share disease loci but have opposite risk alleles ^11^. Given the fact that the alleles of these loci influence expression levels we proposed a theory of trade-offs in aging lung disease ^12^. A trade-off exists whenever a benefit in one context entails a cost in another ^13^. In aging lungs, the high expressing *MUC5B* T-allele may be important for optimal airway defense against infections while it provides an increased risk for IPF in the alveolar compartment.

Therefore, we hypothesize that the high expressing *MUC5B* T-allele of rs35705950 is protective against development of severe COVID-19 disease. We expect that the frequency of the T-allele is significantly lower in patient with COVID-19 requiring hospitalization.

## Material and methods

### Patients

In the study, 108 adult patients hospitalized with COVID-19 at St Antonius Hospital between March 19 2020 and May 5 2020 were included. Diagnosis of COVID-19 were made on the basis of a positive SARS-CoV-2 PCR except for three cases with clinical characteristics and a high-resolution computed tomography (HRCT) congruent with COVID-19 disease. We collected demographics, radiology and survival data from medical hospital records.

The control group consists of 611 Dutch white healthy controls. The study was approved by The Medical research Ethics Committees United (MEC-U) of St. Antonius Hospital and all patients provided written informed consent (approval number R05-08A).

### Genotyping

DNA was extracted from whole blood and genotyped for *MUC5B* rs35705950 genotype with a pre-designed taqman SNP genotyping assay (Applied Biosystems) and the QuantStudio® 5 Real-Time PCR system (ThermoFisher Scientific, Waltham, Massachusetts, USA).

### Statistical analysis

SPSS 24 (IBM, Armonk, New York, USA) was used for statistical analysis. Due to ethnic differences in the prevalence of the *MUC5B* rs35705950 alleles, genetic analyses were stratified by ethnicity and only statistically analyzed in white subjects. Differences between white and non-white patients and between carriers and non-carriers of the rs35705950 T-allele were calculated using a Chi square test for categorical data and Mann-Whitney-U test for non-parametric continuous data and t-test for parametric continuous data. Differences between the allele and genotype frequencies were calculated with the Pearson’s goodness-of-fit chi-square test, together with the OR and 95% CI. Fisher’s exact test was used to test for deviation from Hardy–Weinberg equilibrium. A value of p < 0.05 was considered statistically significant.

## Results

### Patients

In total 108 patients hospitalized with COVID-19 (table 1) were included in the study of which 74 (69%) were males. The median age of the patients was 66 years (range 19.1 – 92.4). Of all patients, 23 patients died (21 %) and they were significantly older than patients who survived, 74 versus 63 years respectively (p=0.002).

**Table 1.**
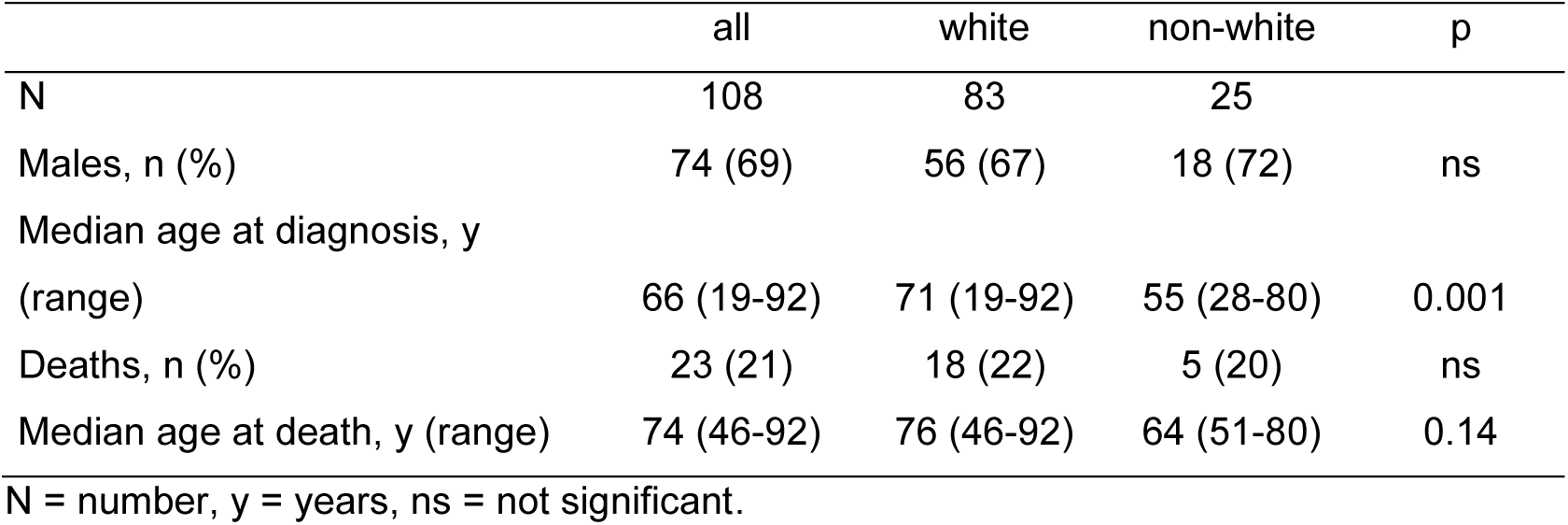
Characteristics of patients hospitalized with COVID-19.

Among 108 patients, 83 (77%) were white and 25 patients were non-whites. The median age at diagnosis differed significantly between whites (71 years) and non-whites (55 years; p=0.0004), and there was a trend towards significance for a younger age at death in non-whites (table 1).

### MUC5B rs35705950

In the total group of 108 patients, there were 99 patients with a GG genotype and 9 patients with a GT genotype. The minor T-allele frequency of the MUC5B promoter polymorphism was 0.04. In the white subgroup of COVID-19 patients, 76 had a GG genotype and 7 a GT genotype, which was in Hardy-Weinberg equilibrium. The frequency of the T-allele in the white COVID-19 group was 0.04 and this was significantly lower than the T-allele frequency of 0.10 in the control group (p=0.023; OR=0.42, CI=0.19-0.91). Moreover, comparison of the number of carriers and non-carriers of the T allele between patients and controls showed that only 8.4% of patients carried the minor allele versus 18% of controls (p=0.029; OR= 0.41, CI=0.19-0.94; table 2).

**Table 2.** *MUC5B* rs35705950 genotype of white subjects with COVID-19 and controls.

|  | COVID-19 | Controls | OR (CI) | p |
| --- | --- | --- | --- | --- |
| N | 83 | 611 |  |  |
| GG, n | 76 | 501 |  |  |
| GT, n | 7 | 103 |  |  |
| TT, n | 0 | 7 |  |  |
| MAF | 0.04 | 0.10 | 0.42 (0.19-0.91) | 0.023 |
| T-carriers, n | 7 | 110 | 0.41 (0.19-0.94) | 0.029 |

N= number, MAF = minor allele frequency, OR = odds ratio, CI = confidence interval.

## Discussion

In this study we examined whether a genetic polymorphism that influences expression of MUC5B is associated with susceptibility to severe COVID-19 disease. We observed a significant association between the *MUC5B* rs35705950 promoter polymorphism and COVID-19 disease. The T-allele frequency and T-carrier frequency was lower in white hospitalized COVID-19 patients than in white controls, suggesting that the T-allele is protective against severe COVID-19 disease.

Beneficial effects of carriership of the T-allele have been reported before. In smoking non-Hispanic white COPD patients with interstitial HRCT features carriers experienced less acute respiratory disease and a longer time-to-first event ^14^. Furthermore, in IPF patients, carriers had a lower bacterial burden than non-carriers ^15^ and better survival ^16^. In congruence with these finding, lower bacterial burden in IPF was associated with less disease progression ^17^.

In the human respiratory system, MUC5B is secreted throughout the lung by submucosal glands and the superficial epithelium of trachea, bronchi, bronchioles and alveoli, and by salivary glands and nasal mucosa^3,5,18,19^. Carriers of the *MUC5B* T-allele demonstrated upregulated RNA expression of MUC5B in lung tissue ^8,9 20^. The increased MUC5B production in T-allele carriers may therefore protect carriers from adverse events related to airway defense. This may be of particular importance in aging subjects, because mucus production and mucociliary clearance have been described to decrease with natural aging ^6,21^. Decreased mucociliary clearance may underlie the observed age-related increase in the incidence of severe community-acquired pneumonia in the elderly ^22^. Similar to previous reports on COVID-19 ^1^ we also observed that severe COVID-19 and death from COVID-19 is predominantly found in the elderly.

With increasing age, the lung changes to the extent that alleles which in younger people confer non-essential divergent expression, may start influencing the risk of disease development in aged tissue. In aging lung diseases such as IPF, COPD and lung cancer, a pattern is emerging of shared disease loci. Interesting however, diseases associate with opposite risk alleles, which oppositely influence expression levels^11,12,23^. Previously we summarized findings and presented a theory in which trade-offs between compartments of the aging respiratory system exist, with the different compartments requiring different optimal expression levels ^12^.

The MUC5B rs35705950 polymorphism may be added to this list of shared loci, because the T-allele, which appears beneficial in this and above described studies, is best known as a major risk allele for IPF ^8^. IPF is a rare non-infectious pulmonary aging disease of unknown cause characterized by insidious onset of disease in patients without a history of pulmonary infection. Subsequent research showed that the MUC5B T-allele not only predisposes to IPF but to a variety of chronic progressive forms of pulmonary fibrosis ^24-27^. The key cell in the pathogenesis of pulmonary fibrosis is the alveolar type II (AT2) cell. Recent research showed that ectopic overproduction of Muc5b in AT2 cells in mice cause mucociliary dysfunction and bleomycin-induced pulmonary fibrosis ^19^. Interestingly, overproduction in AT2 cells caused worse mucociliary dysfunction than overproduction in club cells in airways ^19^.

The fact that the high expressing T allele is associated with an increased risk for pulmonary fibrosis combined with the current and previous findings that the T-allele protects against severe COVID-19 and infectious burden is another example of a trade-off that arises with aging. During the first decades of life the effect of both alleles is neutral while at an older age differences in constitutive expression predispose to disease. The price for enhanced expression in the airways and better defense against viral pathogens may be early aging of the alveolar compartment.

Although the finding is of interest it must be noted that the small sample size and the lack of replication is a major limitation of the current study. Furthermore, due to limited sample size we were not able to perform an association analysis in non-white subjects, while they suffer disproportionally from COVID-19. In conclusion, we found that the T-allele of MUC5B rs35705950 confers protection from development of severe COVID-19 disease. Because the T-allele is a known risk allele for pulmonary fibrosis, the study provides further evidence for the existence of trade-offs between optimal expression levels in the aging lung. Further studies with lager sample size are needed to understand the importance of our finding.

## Data Availability

Data may be requested from the corresponding author

## Acknowledgement

We thank Mirjam Visser for support in the patient informed consent procedure.

## Funding

ZonMW TopZorg St Antonius Care grant nr 842002001; ZonMW Topspecialistische Zorg en Onderzoek grant nr 10070012010004; Nederlandse Vereniging van Artsen voor Longziekten en Tuberculose COVID-19 grant

## Notes

### Competing Interest Statement

The authors have declared no competing interest.

